# Circulating Tumor DNA As A MRD Assessment And Recurrence Risk In Patients Undergoing Curative Intent Resection With Or Without Adjuvant Chemotherapy In Colorectal Cancer: A Meta-analysis

**DOI:** 10.1101/2022.11.04.22281967

**Authors:** Anusha Chidharla, Eliot Rapoport, Kriti Agarwal, Samragnyi Madala, Brenda Linares, Weijing Sun, Sakti Chakrabarti, Anup Kasi

**Author notes:** Contributed equally as first authors (Anusha Chidharla, Eliot Rapoport).

## Abstract

**PURPOSE:** Emerging data have suggested that circulating tumor DNA (ctDNA) can be a reliable biomarker for Minimal residual disease (MRD) in CRC patients. Recent studies have shown that the ability to detect MRD using ctDNA assay after curative-intent surgery will change how to assess recurrence risk and patient selection for adjuvant chemotherapy.

**METHODS:** We performed a meta-analysis of post-operative ctDNA in Stage I-IV (oligometastatic) CRC patients after curative-intent resection. We included 23 studies representing 3,568 patients with evaluable ctDNA in CRC patients post-curative intent surgery. Data were extracted from each study to perform a meta-analysis using RevMan 5.4. software. Subsequent subgroup analysis was performed for stages I-III and oligometastatic stage IV CRC patients.

**RESULTS:** The pooled hazard ratio (HR) for recurrence-free survival (RFS) in post-surgical ctDNA positive versus negative patients in all stages was 7.27 (95% CI 5.49-9.62) p <0.00001. Subgroup analysis revealed pooled HR of 8.14 (95% CI 5.60-11.82) and 4.83 (95% CI 3.64-6.39) for stage I-III and IV CRC, respectively. The pooled HR for RFS in post-adjuvant chemotherapy ctDNA positive versus negative patients in all stages was 10.59 (95% CI 5.59-20.06) p <0.00001. The subgroup analysis based on the ctDNA method showed a pooled HR of 8.66 (95% CI 6.38-11.75) and 3.76 (95% CI 2.58-5.48) for tumor-informed and tumor-agnostic, respectively.

**CONCLUSION:** Our analysis emphasizes that post-operative ctDNA is a strong prognostic marker of RFS. Based on our results, ctDNA can be a significant and independent predictor of RFS. This real-time assessment of treatment benefits using ctDNA can be used as a surrogate endpoint for the development of novel drugs in the adjuvant setting.

## Introduction

Cell-free DNA (cfDNA) are small DNA fragments (160-200 bp) released into the bloodstream during cell death. In healthy adults, cfDNA is primarily released by hematopoietic cells; however, in the setting of cancer, many tumors also release DNA fragments, referred to as circulating tumor DNA (ctDNA), into the systemic circulation. ^1–3^ CtDNA has a short half-life of approximately 2 hours. This property allows it to be used as a dynamic marker for tracking the presence of the tumor. ^4,5^ Although somewhat limited by the delayed turnaround time and cost, there is significant interest in ctDNA. It is a minimally invasive test, which, given its dynamic nature, has high sensitivity and specificity. ^1,6,7^

Two forms of ctDNA analysis have been developed: tumor-informed techniques and tumor-agnostic or tumor-naive techniques. In tumor-informed methods (e.g., signatera and SafeSeqS), first somatic mutations are first identified in tumor tissue, followed by targeted sequencing of plasma DNA using a personalized assay. Tumor-informed assays have better sensitivity and specificity than tumor-agnostic assays where generic gene panels are used (e.g., guardant).^3,8,9^ A significant drawback is the prolonged turnaround time required for personalization. Both methods are currently being evaluated despite cost concerns, hematopoiesis-associated false positives, and reproducibility.

The utility of ctDNA is being explored in numerous contexts, with evidence supporting its role in early cancer detection, monitoring treatment response, and evaluating recurrence and efficacy for multiple forms of cancer. One specific area of interest is its role in assessing minimal residual disease (MRD) and the possibility of its use to guide therapeutic decisions. One hope is that it will be able to guide treatment in the controversial setting of adjuvant chemotherapy (ACT) in stage II and other non or oligometastatic colorectal cancers (CRC). The role of ACT in this setting is poorly defined because of the heterogeneity within disease stages. ^10^

The benefit of adjuvant 5-FU-based chemotherapy in locally advanced colon cancer has been recognized since the late 1980s. A meta-analysis published by Buyse et al. in 1988 comparing adjuvant 5-FU with surgery alone favored adjuvant chemotherapy with a mortality odds ratio of 0.83 (95% CI 0.70-0.98.^11^ This was established by North Central Cancer Therapy Group (NCCTG)2 and Intergroup (INT)-00353 trials, which formed the basis for current guideline recommendations to include 5-FU-based adjuvant chemotherapy in stage II/III colon cancer patients. While the guidelines for adjuvant chemotherapy in stage III colon cancer are unambiguous, its use in stage II disease is debatable—especially considering the toxicity associated with chemotherapy regimens with unclear benefits. ^11,12^Current guidelines recommend 3-6 months of adjuvant chemotherapy after surgery for nonmetastatic colon cancer.^13^ In patients deemed to have high-risk Stage II CRC, surgery is followed by adjuvant chemotherapy. This decision is made based on tumor size as well as pathological and clinical features of the disease, which are relatively poor predictors.^13^ Not all patients require ACT, and it has been challenging to determine what subset does.^10,11,14,15^ Henceforth, there is a need for predictive and prognostic biomarkers for follow-up detection of early recurrence, thereby enabling appropriate follow-up and therapeutic strategies for early recurrence detection and curative treatment.

Recent advances in technology in ctDNA assay can detect minimal residual disease (MRD) after curative intent surgery. ^16,17^Using ctDNA to guide the treatment can help avoid the toxic effects of chemotherapy after surgery, especially in patients with a low risk of recurrence. ctDNA has been shown to have a prognostic value and is a good predictor of cancer recurrence in many recent studies. ^18^Emerging data have suggested that circulating tumor DNA (ctDNA) can be a reliable biomarker for MRD. This may change how to assess recurrence risk and patient selection for adjuvant chemotherapy. ^19^ Therefore, we conducted a systematic review and meta-analysis of studies evaluating the value of ctDNA in the post-surgical and post-ACT periods to predict prognosis and recurrence.

## Materials and Methods

This systematic review and meta-analysis were exempt from institutional review board approval based on Kansas University Medical Center criteria. The study was conducted in accordance with the Preferred Reporting Items for Systematic Reviews and Meta-analyses (<u>PRISMA</u>) recommendations.

A professional librarian searched PubMed/Medline, EMBASE Web of Science, Cochrane Library, and Google from the database inception through June 8, 2022, using Keywords, Medical Subject Heading (MeSH), and EMTREE subject headings used to search for the concepts of colon cancer, ctDNA, survival, and types of studies. The search included full-text articles and conference presentations. The search terms *colorectal neoplasm* AND *circulating tumor DNA* were expanded and used with appropriate MeSH terms. The results were refined according to the study type and outcomes.

### Study Eligibility

Studies were evaluated by at least two independent reviewers (AC, ER, KA), with a third confirming the final inclusion and resolving disagreements (AK). Studies were chosen on the basis of the following criteria: (1) randomized clinical trials or prospective/retrospective cohort studies, (2) patients with stage I-III or oligometastatic stage IV colorectal cancer; (3) studies examining post-operative ctDNA status or post-adjuvant ctDNA status (4) ctDNA data were derived from a panel of mutations rather than single mutations (5) data were available on patient outcomes including disease-free survival, recurrence-free survival, or overall survival (6) the data is not better represented in another entry; (7) both full published manuscripts and conference abstracts were included. Studies beyond the inclusion criteria or those originally published in a language other than English were excluded.

### Data Extraction

Extraction was performed by at least two reviewers (AC, ER, KA), with disputes resolved by discussion with the third. Data were recorded regarding study characteristics, patient demographics, stages studied, ctDNA collection method, the timing of ctDNA collection, and reported Recurrence Free Survival (RFS)/ Recurrence Free Interval (RFI) in both post-surgical and post-adjuvant chemotherapy periods. In addition, data were recorded for individual subgroups such as stages and the study at large when available.

### Statistical Analysis

Data analysis was performed using Review Manager V.5.3 (The Nordic Cochrane Center, Cochrane Collaboration, Copenhagen, Denmark). If the study had more than one outcome, precision was compared to give a more conservative estimate of the HRs and 95%□CI. The I^2^ statistic was used to assess the statistical heterogeneity. An I^2^ statistic of >50% was considered significant heterogeneity. Statistical significance was set at p value <0.05. Publication bias was assessed visually using funnel plots. All studies were assessed to be of moderate quality. The pooled HR and 95%□CI are represented in forest plots. Each square on the chart area represents an individual study, and the area of each square is equivalent to the weight of the study, which is the inverse of the study variance. The diamond represents summary measures, and the width corresponds to the 95%□CI. A random-effects model with inverse variance (DerSimonian and Laird method) was applied.^20^ Heterogeneity was estimated using the inconsistency index and χ^2^ test.

## Results

Our search yielded a total of 668 articles. After screening and final selection, 23 unique studies provided quantitative data on RFS based on the post-operative and post adjuvant ctDNA status. A PRISMA diagram is shown in Figure 1. The characteristics of these studies are summarized in Table 1. Henricksen et al. 2021 and Henricksen et al. 2022 were duplicates but were used for different analyses.^18,19^ Of these studies, seven provided data on the prognostic value of post-adjuvant ctDNA. The studies primarily focused on locally invasive or otherwise nonmetastatic cancers, although eight studied ctDNA in oligometastatic stage IV rectal cancer amenable to curative intent resection. Most studies (17/23) utilized a tumor-informed ctDNA analysis method.

**Table 1:**
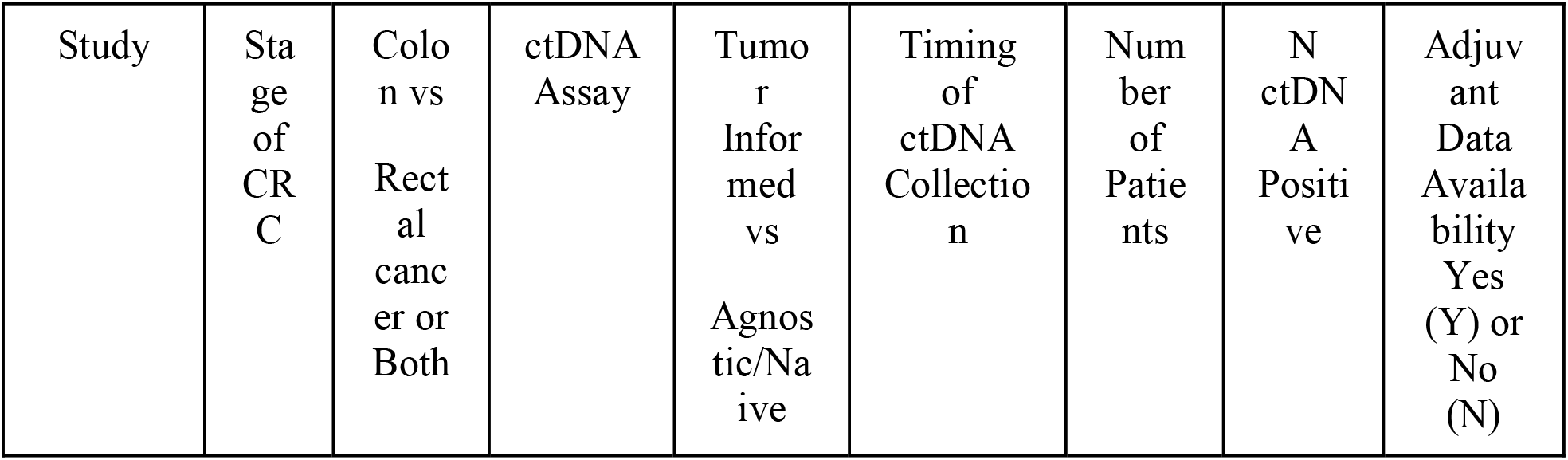
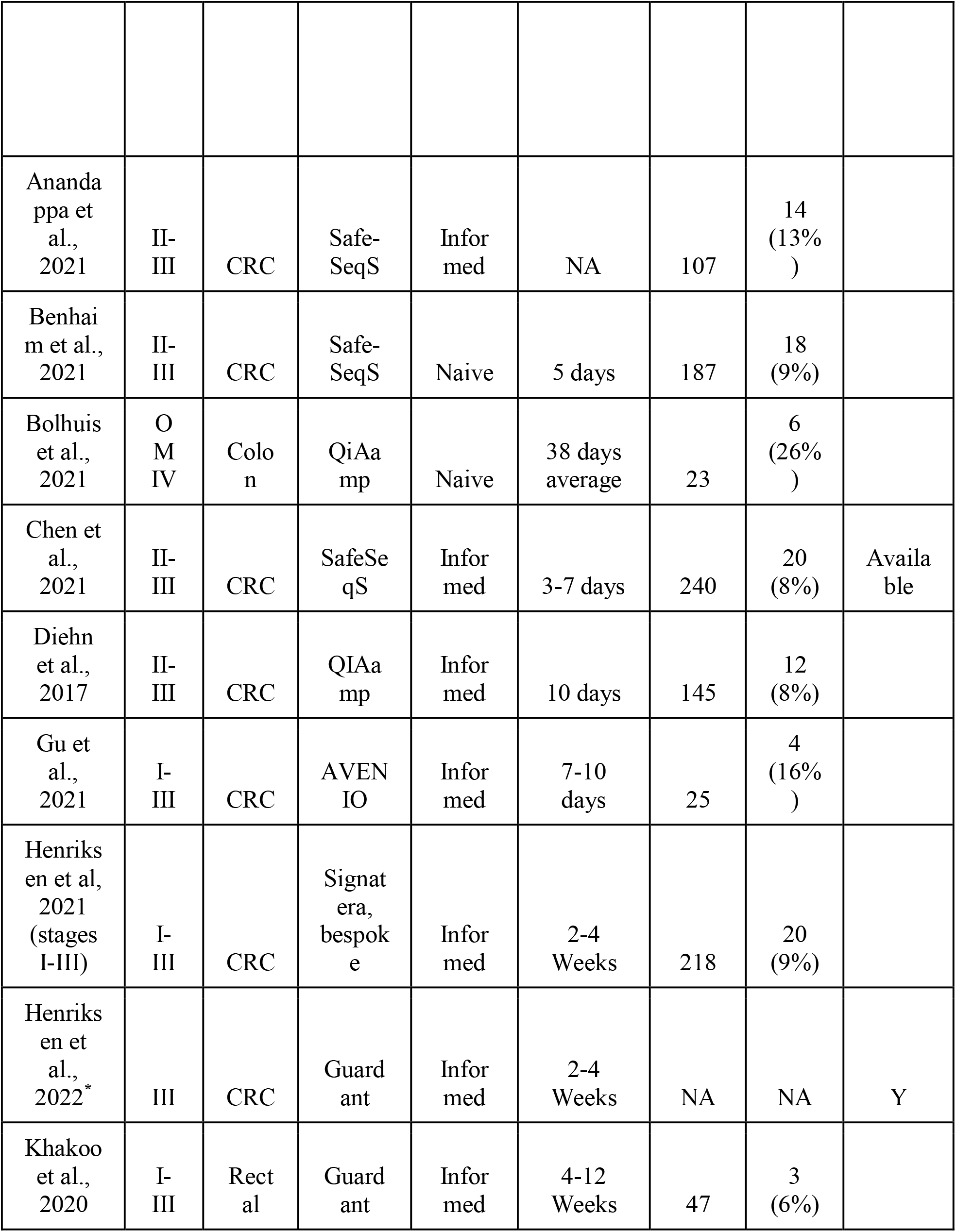
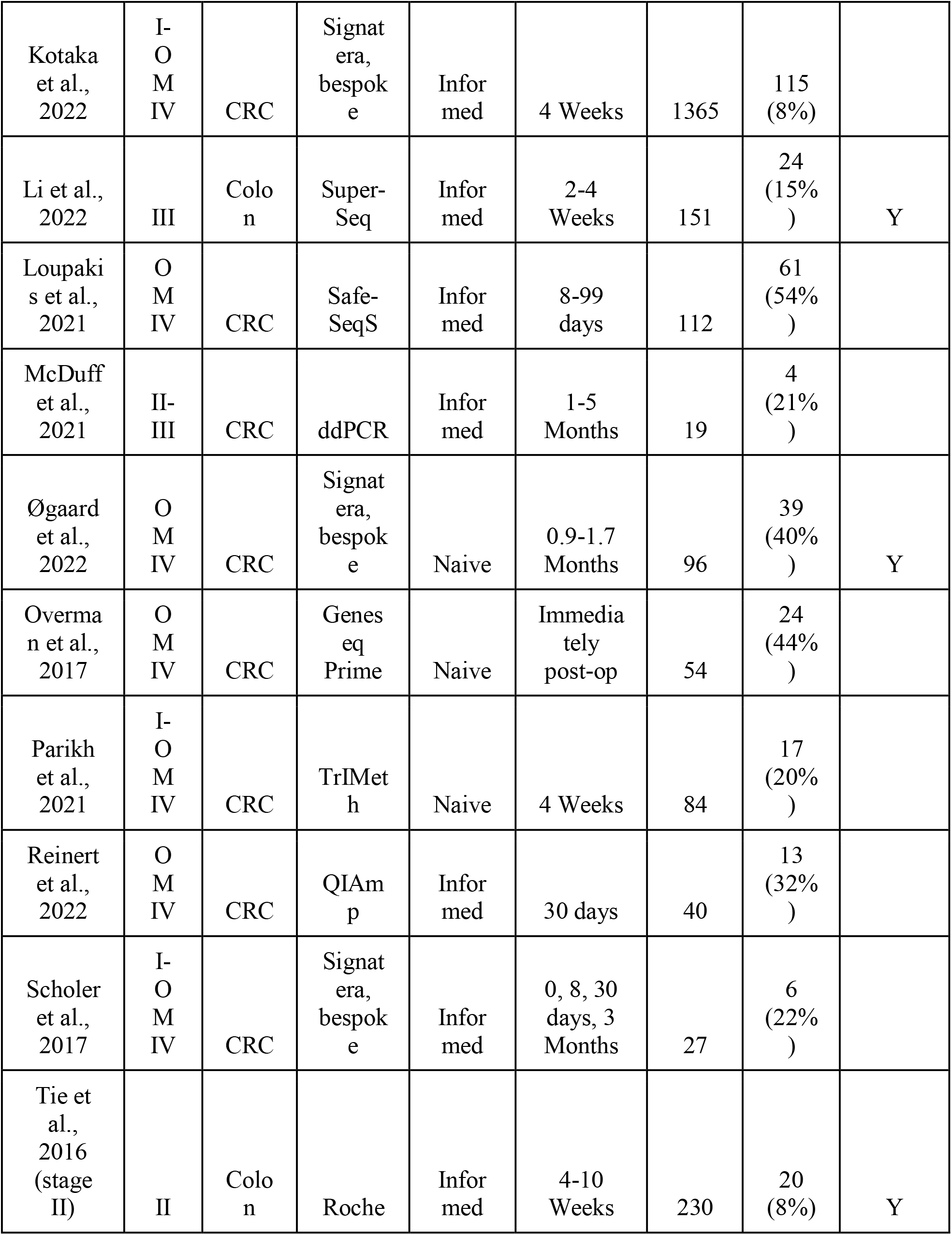
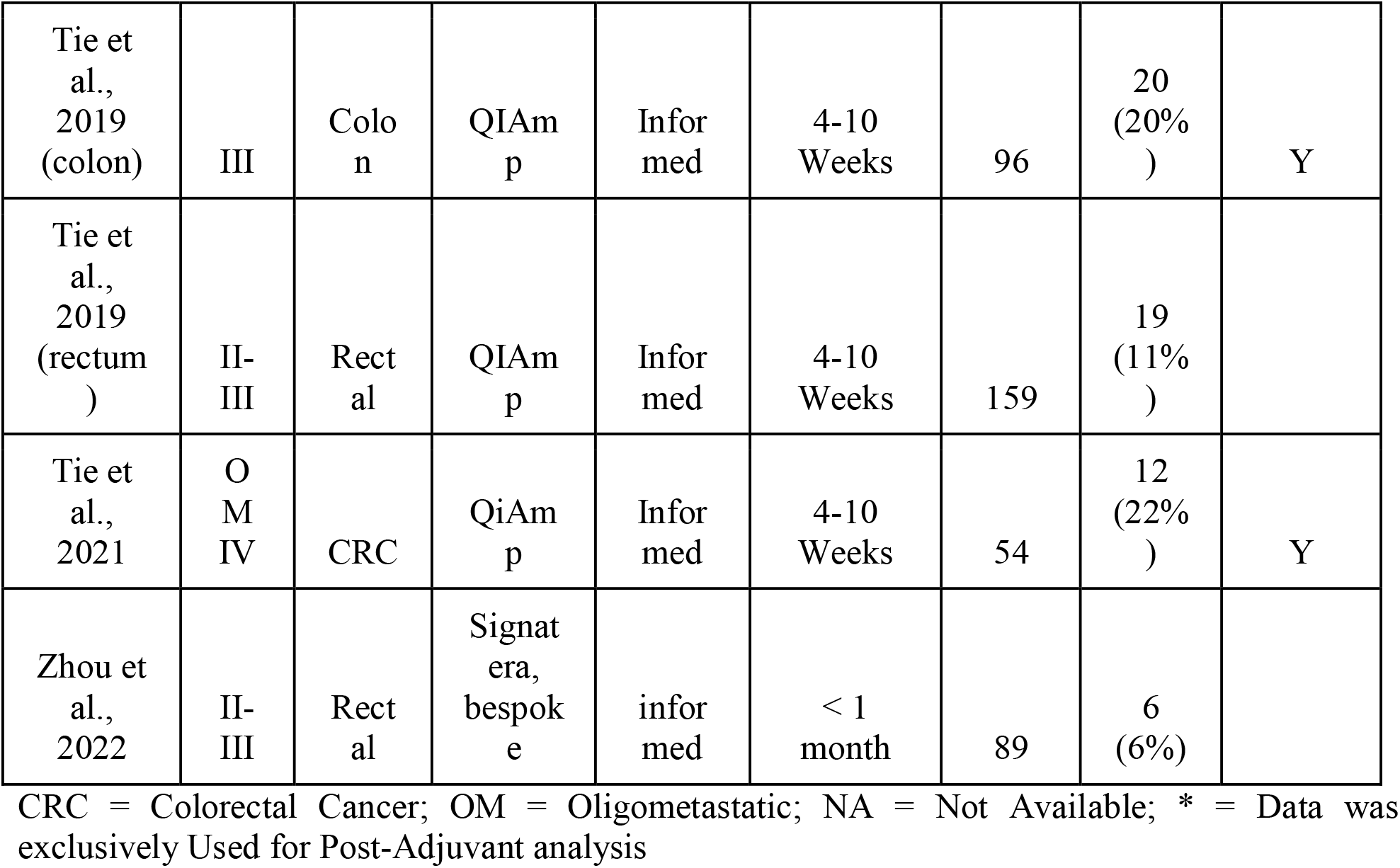
Characteristics of the studies included in this meta-analysis.

**Figure 1:**
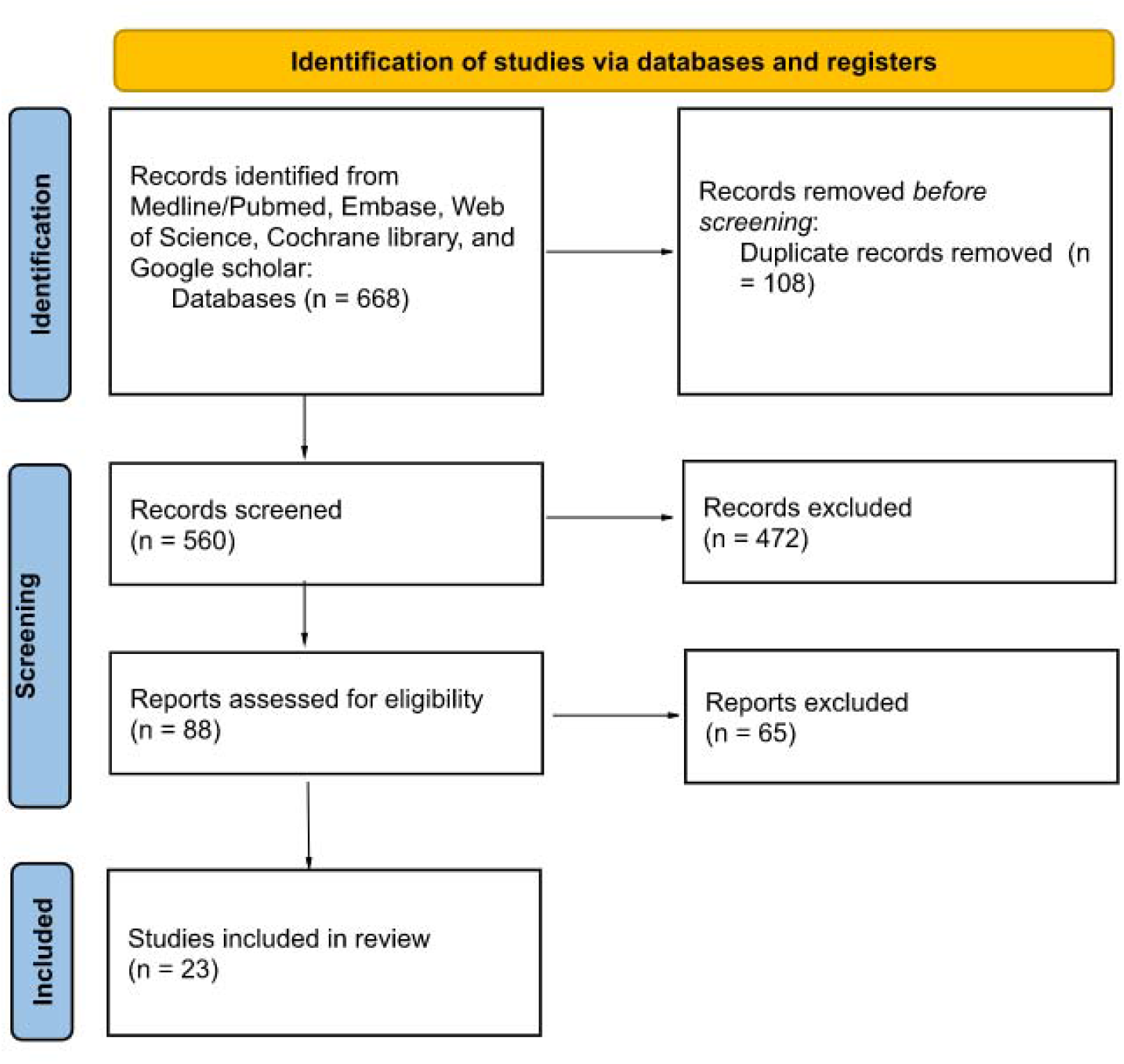
PRISMA flow diagram

The data comprised of 3,568 patients. Of this population, 13.4% (477) were positive for ctDNA post-operatively. Likewise, 1,007 patients were assessed in the post-adjuvant setting.

Utilizing a random-effects model, analysis of our primary outcome, post-surgical ctDNA status, showed a statistically significant prognostic effect, pooled HR= 7.27 (95% CI 5.49-9.62, P < 0.0001). This indicates that the presence of a positive ctDNA results after surgery yields a poor prognosis. A forest plot of this data is shown in Figure 2. These data had moderate heterogeneity (I^2^=55%). Subgroup analyses were performed on these data, as shown in Table 2. There have been insufficient studies to stratify post-adjuvant ctDNA results for oligometastatic stage IV disease and tumor-agnostic methodologies. Among these analyses, all the pooled hazard ratios reached significance. Heterogeneity was improved when stratifying by the tumor-informed versus tumor-agnostic ctDNA collection method, especially in the tumor-informed group. Similarly, heterogeneity improved when only oligometastatic stage IV was analyzed. Forest plots of tumor agnostic and tumor-informed ctDNA statuses are shown in Figure 3.

**Table 2:** Pooled Hazard Ratio for Subgroup Analyses based on Stage, Method of ctDNA analysis.

| Subgroup | Pooled HR (CI) | Number of Studies |
| --- | --- | --- |
| **Post-surgical** | **7.27 (95% CI 5.49-9.62)** | **22** |
| Stage |  |  |
| *I-III* | *8.14 (95% CI 5.60-11.82)* | *14* |
| *IV oligometastatic* | *4.83 (95% CI 3.64-6.39)* | *8* |
| ctDNA Method |  |  |
| *Tumor-Informed* | 8.66 (95% CI 6.38-11.75) | 17 |
| *Tumor-Agnostic* | 3.76 (95% CI 2.58-5.48) | 5 |
| **Post-Adjuvant** | 10.59 (95% CI 5.59-20.06) | 7 |
| Stage |  |  |
| *I-III* | 10.60 (95% CI 4.21-26.69) | 5 |
| *IV oligometastatic* | NA | 2 |
| ctDNA Method |  |  |
| *Tumor-Informed* | 11.16 (95% CI 5.19-23.98) | 6 |
| *Tumor-Agnostic* | NA | 1 |

**Figure 2:**
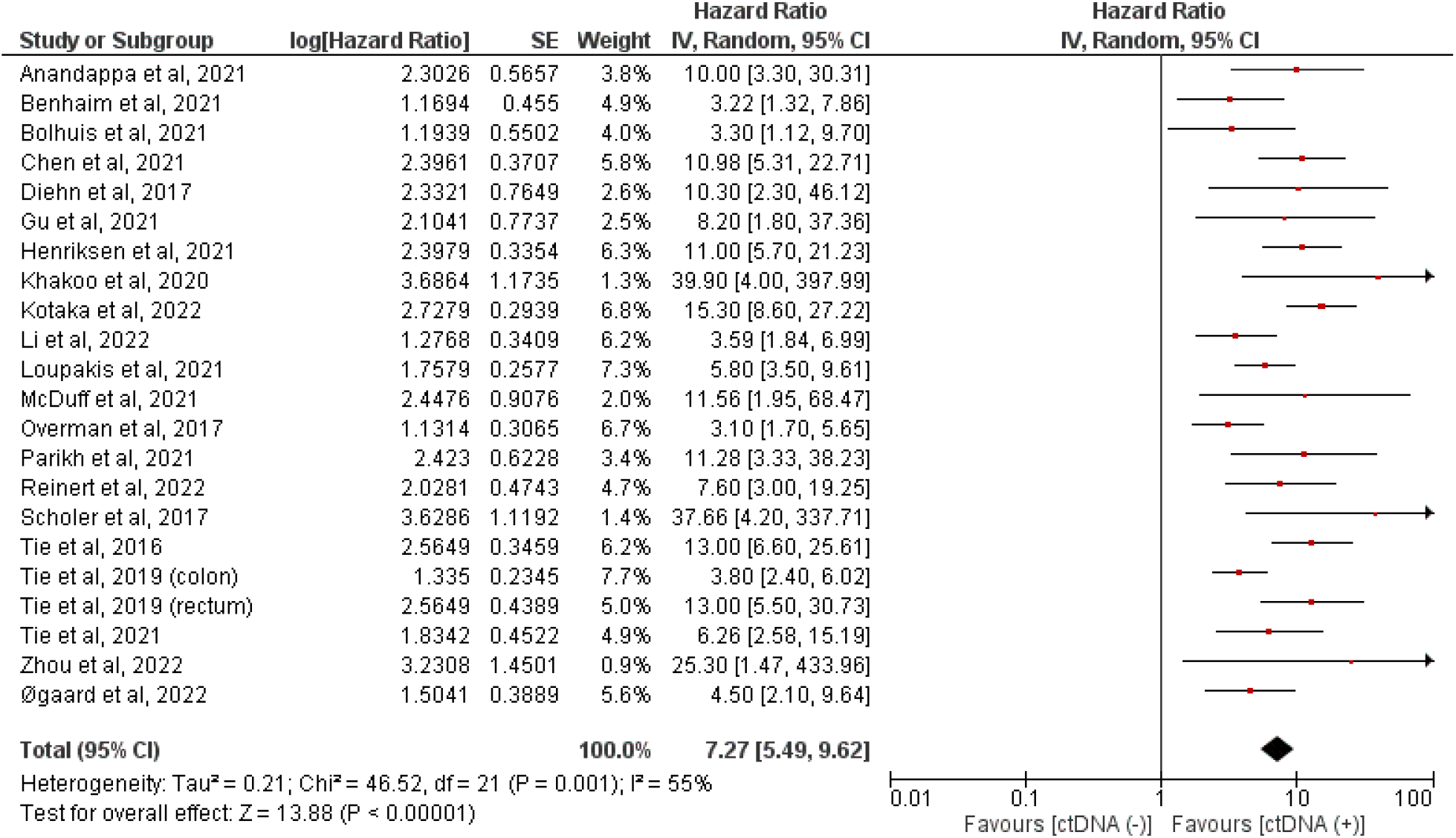
Forest plot showing the pooled hazard ratio based on post-surgical ctDNA positive versus ctDNA negative status. The hazard ratio for each adverse event is represented by a square, and the horizontal lines crossing the squares represent the 95% confidence interval (CI).

**Figure 3:**
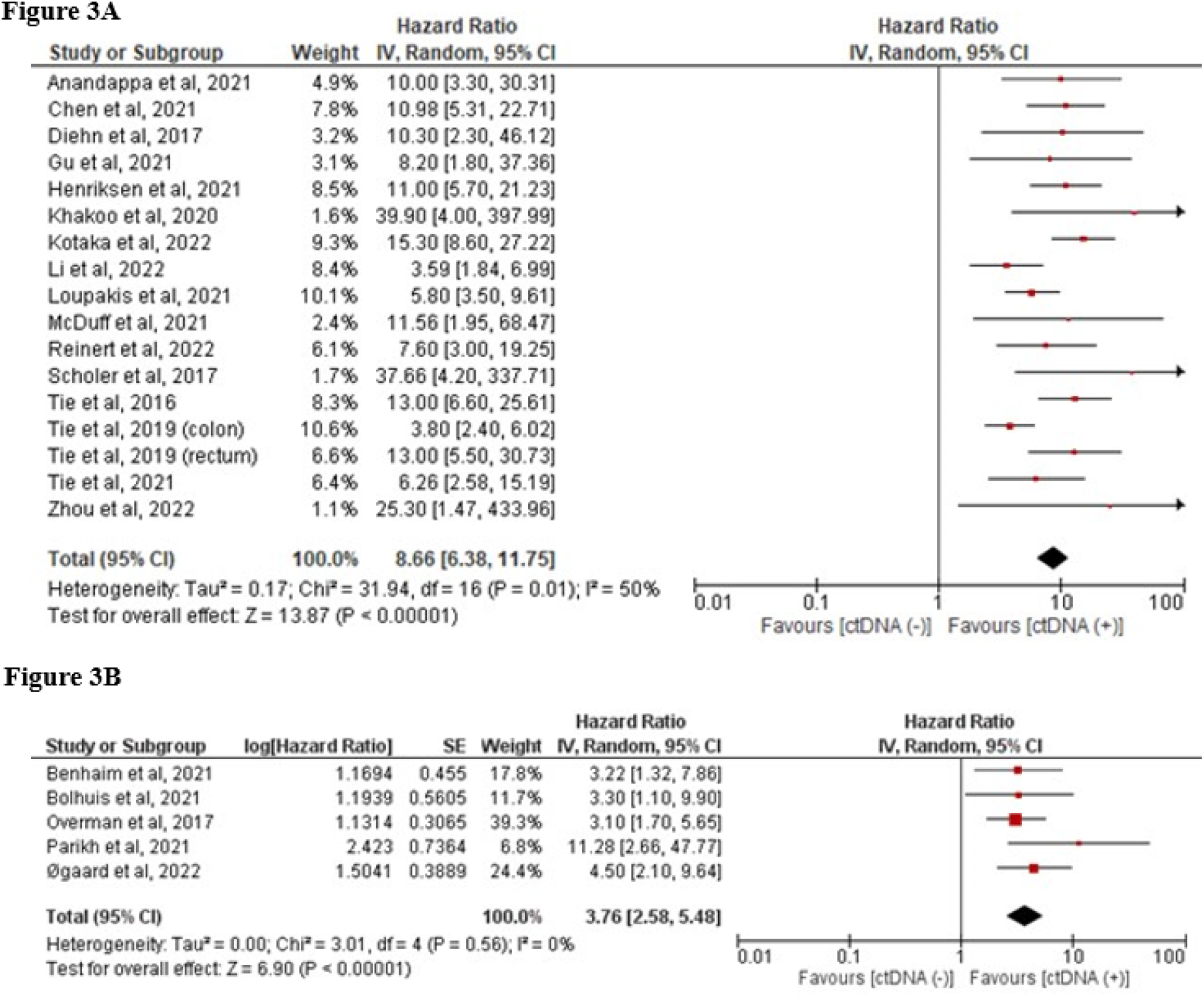
Forest plot showing the pooled hazard ratio based on ctDNA method (A) post-surgical ctDNA positive versus ctDNA negative status via the tumor-informed method. (B) post-surgical ctDNA positive versus ctDNA negative status via tumor agnostic method. The hazard ratio for each adverse event is represented by a square, and the horizontal lines crossing the squares represent the 95% confidence interval (CI).

Similarly, a random-effects model was used to calculate the pooled HR for ctDNA status in the post-adjuvant chemotherapy setting, which also yielded a statistically significant result that positive ctDNA implies a higher risk of recurrence, pooled HR = 10.59 (95% CI 5.59-20.06) (Figure 4). Unfortunately, a meta-analysis could only be performed on stages I-III and tumor-informed methodology in the post-adjuvant setting owing to the smaller number of studies.

**Figure 4:**
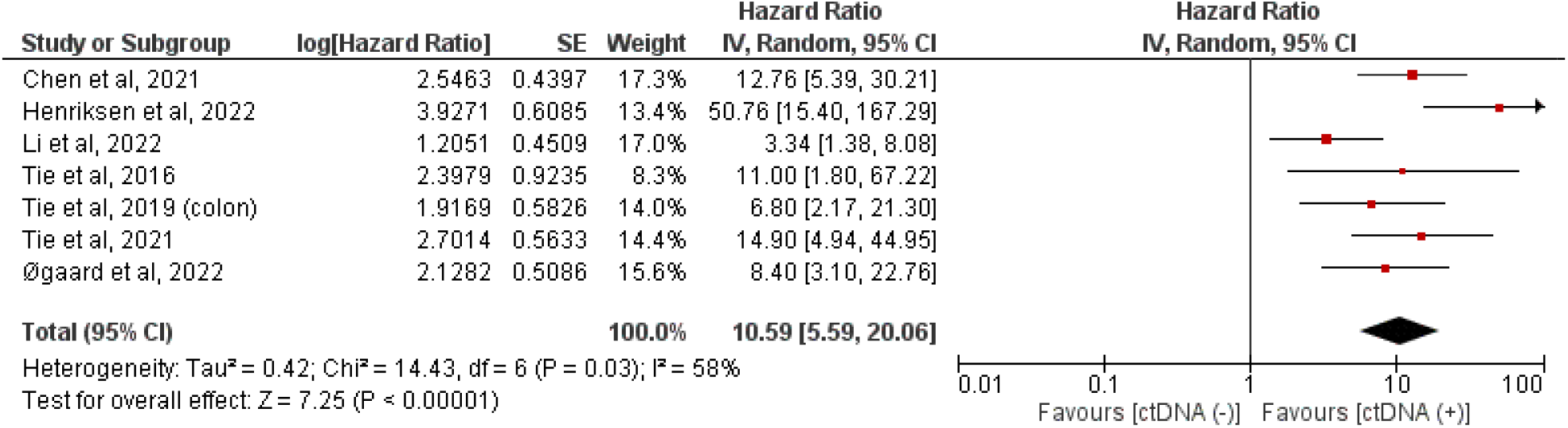
Forest plot showing the pooled hazard ratio based on the ctDNA method based on post-adjuvant ctDNA positive versus ctDNA negative status. The hazard ratio for each adverse event is represented by a square, and the horizontal lines crossing the squares represent the 95% confidence interval (CI).

## Discussion

Our study demonstrated that patients with ctDNA-positive status after curative intent surgery was significantly associated with low RFS, pooled HR= 7.27 (95% CI 5.49-9.62, p < 0.0001). This indicates that patients with positive ctDNA following curative-intent surgery have a poorer prognosis than ctDNA-negative patients. Based on these results, ctDNA analysis can reliably identify patients at a higher risk of recurrence and those who can benefit from adjuvant systemic treatments. This could spare patients from unnecessary or inappropriate toxic treatments. Therefore, ctDNA analysis could also be used as a predictive marker. A phase II/III study, NRG-GI005 (COBRA), is currently testing whether ctDNA can be a predictive biomarker for adjuvant chemotherapy benefit in patients with resected Stage II colon cancer. ^21^

After practicing for decades with no reliable minimally invasive marker, it is practice changing to now have post-operative ctDNA analysis to help guide our decisions regarding adjuvant therapy. Our study is the largest meta-analysis to explore the role of ctDNA assay. A smaller meta-analysis of 7 studies with data prior to 2019 included 424 patients and showed a statistically significant association between post-surgical ctDNA and RFS.^22^ The current prospective studies with ctDNA have a small number of patients and do not reflect the true value of ctDNA in MRD monitoring. ^23^ Our meta-analyses included 23 studies with 3,568 patient ^16,18,19,24–32^. This speaks to the rapidly expanding number of studies on the topic. Synthesizing an emerging abundance of robust data is essential.

We performed subgroup analyses of patients based on the CRC stage. Patients with stage I-III CRC are eight times more likely to recur with positive ctDNA results than ctDNA negative patients. This provides an indicator for patients who may benefit from further adjuvant treatment to prevent recurrence. Further studies on this topic are ongoing. CIRCULATE-Japan, which encompasses three clinical trials, is currently examining the clinical benefits of ctDNA analysis and adjuvant treatment in patients with resectable CRC. ^33,34^

Our analysis also showed that patients who had positive ctDNA after receiving adjuvant chemotherapy had a poorer prognosis with lower RFS than ctDNA-negative comparators (HR 10.59, 95% CI 5.59-20.06). Post-adjuvant ctDNA levels could be used to determine the risk of recurrence and the need for further close surveillance.^35,36^ For example, nearly half of the patients with stage IV CRC with liver oligometastases recur after curative intent surgery. Reinert and colleagues studied these patients with serial ctDNA studies in addition to routine surveillance imaging.^37^ The study showed that ctDNA detected recurrence with a median time of 2.5 months (p< .0001) prior to routine surveillance imaging, especially in those with indeterminate CT findings. This indicates that ctDNA can be used as a surveillance tool to assess recurrence. In a study of 138 patients with metastatic gastrointestinal cancer, Parikh et al. found that serial ctDNA monitoring could predict the response to systemic treatment. ^38^Currently NRG-GI008 trial is recruiting patients with Stage III and high-risk Stage II colon cancer to determine which patients benefit from adjuvant chemotherapy based on the ctDNA results. ^39^

We also explored the role of different ctDNA analysis methods on prognostications, as previous studies have shown that tumor-informed methods are more sensitive and specific compared to tumor-agnostic methods. We performed a subgroup analysis of ctDNA analysis methods in the post-surgical setting. This demonstrated that ctDNA positivity using tumor-informed and tumor-agnostic methods was associated with low RFS with pooled hazard ratios of 8.66 (95% CI 6.38-11.75) and HR=3.76 (95% CI 2.58-5.48), respectively.^40,41^ While this data indicates better prognostication for tumor-informed methodologies in line with previously published information, and the studies included did not include head-to-head analysis, so conclusive arguments are difficult to make from this data. However, this is consistent with previously published results, which showed that studies that used tumor-informed assays showed higher rates of recurrences than those that used tumor-agnostic assays.

Monitoring ctDNA levels in the blood has been shown to accurately detect MRD and aid in measuring the therapeutic effects after curative treatment. While ctDNA is not yet the standard of care in clinical practice for CRC patients, studies are ongoing to define the appropriate way to use it as a tool in the clinic. ^42–44^ In 2022, a phase two randomized trial, Circulating Tumor DNA Analysis Informing Adjuvant Chemotherapy (DYNAMIC), showed non-inferiority in 2-year recurrence-free survival between the standard management group and ctDNA-guided management in stage II CRC patients after curative intent surgery (93.5% vs. 92.4%, 95% CI [-4.1 to 6.2], non-inferiority margin, -8.5 percentage points). ^45^ Our study builds on this issue. To our knowledge, this is the largest meta-analysis to confirm the prognostic and predictive power of ctDNA levels in the post-operative and post-ACT settings.

### Limitations

Abstracts with insufficient or imprecise data were excluded. Studies that included Stage 0 CRC were omitted if they lacked subgroup analysis excluding this population. The sensitivity and specificity of the ctDNA methods are different. We attempted to overcome this by performing subgroup analysis of tumor-naive vs. tumor-informed techniques. Multiple abstracts were published on the same population at different time points during follow-up, so we eliminated the duplicates by reviewing all abstracts and manuscripts in detail and included only the most recent abstract or manuscripts with the greatest patient population. As this is a new technique, many studies have lacked extensive follow-up. There are also limited studies for certain populations that prevent meta-analyses from being performed. Additionally, Oligometastatic CRC patients were found to have a lower hazard ratio than earlier-stage cancers, which could be secondary to the limited number of studies in this setting.

## Conclusion

Our study is the largest and most up-to-date meta-analysis of studying the effect of ctDNA status in both post-curative intent surgery and post-ACT in CRC. Our study validated the role of ctDNA analysis in stage I to oligometastatic stage IV CRC. Our analysis emphasizes that post-operative ctDNA is a strong prognostic marker of RFS. Based on our results, ctDNA can be a significant and independent predictor of RFS. This real-time assessment of treatment benefits can be used as a surrogate endpoint for the development of novel drugs. Few ctDNA-based clinical trials are ongoing internationally to confirm the clinical utility of ctDNA in colorectal cancer. Further randomized clinical trials, in which ctDNA results are used to inform patient management, are required to assess the clinical utility of ctDNA-guided approaches for colorectal cancer management and surveillance.

## Data Availability

All data produced in the present study are available upon reasonable request to the authors.
All data produced in the present work are contained in the manuscript.

